# Disruptive Mood Dysregulation Disorder: symptomatic and syndromic thresholds and diagnostic operationalization

**DOI:** 10.1101/19002436

**Authors:** Paola Paganella Laporte, Alicia Matijasevich, Tiago N. Munhoz, Iná S. Santos, Aluísio J. D. Barros, Daniel Samuel Pine, Luis Augusto Rohde, Ellen Leibenluft, Giovanni Abrahão Salum

**Author notes:** **Address correspondence and reprint requests** Paola Paganella Laporte, Hospital de Clínicas de Porto Alegre, Ramiro Barcelos, 2350 – room 2202; Porto Alegre, Brazil – 90035-003.

## Abstract

**Objective:** The aim of this study is to identify the most appropriate threshold for Disruptive Mood Dysregulation Disorder (DMDD) diagnosis and the impact of potential changes in diagnostic rules on prevalence levels in the community.

**Method:** Trained psychologists evaluated 3,562 pre-adolescents/early adolescents from the 2004 Pelotas Birth Cohort with the Development and Well-Being Behavior Assessment (DAWBA). The clinical threshold was assessed in three stages: symptomatic, syndromic and clinical operationalization. The symptomatic threshold identified the response category in each DAWBA item which separates normative misbehavior from a clinical indicator. The syndromic threshold identified the number of irritable mood and outbursts needed to capture pre-adolescents/early adolescents with high symptom levels. Clinical operationalization compared the impact of AND/OR rules for combining irritable mood and outbursts on impairment and levels of psychopathology.

**Results:** At the symptomatic threshold, most irritable mood items were normative in their lowest response categories and clinically significant in their highest response categories. For outbursts some indicated a symptom even when present at only a mild level, while others did not indicate symptoms at any level. At the syndromic level, a combination of 2 out of 7 irritable mood and 3 out of 8 outburst indicators accurately captured a cluster of individuals with high level of symptoms. Analysis combining irritable mood and outbursts delineated non-overlapping aspects of DMDD, providing support for the OR rule in clinical operationalization. The best DMDD criteria resulted in a prevalence of 3%.

**Conclusion:** Results provide information for initiatives aiming to provide data-driven and clinically oriented operationalized criteria for DMDD.

## Introduction

Temper outbursts and irritable mood are common manifestations of typical development. When outbursts and irritable mood are intense, frequent, last for significant periods, occur in several contexts, and are associated with behaviors not seen in typically developing children, they often require clinical attention ^1–3^. Disruptive Mood Dysregulation Disorder (DMDD) is a new diagnosis designed to capture pathological manifestations of irritable mood and temper outbursts ^4^. Given the newness of DMDD, data-driven approaches based on epidemiological evidence are needed to evaluate appropriate thresholds for DMDD and consider the need to refine criteria. The current report provides such data.

DMDD has its origins in the mid-2000s when Leibenluft and colleagues ^5,6^ defined a syndrome called severe mood dysregulation (SMD). SMD involved severe, chronic grouchy mood and heightened reactivity, along with symptoms of hyperarousal ^6^. The syndrome was defined to distinguish children with severe irritability from those with classic bipolar disorder (BD), in light of increasing numbers of children diagnosed with BD ^7,8^. The results of those studies converged to differentiate SMD from classic bipolar disorder based on course and familial aggregation ^9–11^. For DSM-5, SMD was modified to create DMDD.

Alternative thresholds for defining DMDD have been only partially considered in the current literature. Some previous studies have focused on irritability as a dimensional trait, which is broader than DMDD as a diagnostic entity. These studies provide an important framework for investigating clinically-relevant thresholds for specific behaviors. Wakschlag and collaborators ^12^ used item response theory analysis to disentangle normative misbehavior from clinically significant problems by studying the ‘symptomatic threshold’, i.e., investigating which response category in each item from a questionnaire separates normative misbehavior from a clinical indicator. They found that some behaviors are normative and only represent problems when their frequency is high or very high, whereas others always indicate a significant problem that requires clinical attention. This and similar research efforts in preschoolers ^13^ inform attempts to evaluate varying boundaries for the definition of DMDD. Other studies focused more specifically on varying DSM-5 criteria for DMDD in pre-adolescents ^14,15^ and adolescents ^14–16^. They found the prevalence of temper outbursts and negative mood are much lower than what is found in preschoolers and that applying exclusion criteria such as frequency and hierarchical diagnostic rules affects DMDD prevalence rates considerably ^14–16^. There was no evidence that clinical markers changed between pre-adolescents/early adolescents (9-12) to middle adolescents (13-16) ^15^. Nonetheless, it is important to continue to identify appropriate diagnostic thresholds for distinct developmental periods, given that normative levels of irritability clearly vary across the lifespan ^14–16^.

Another important step towards evaluating such varying boundaries involves quantifying the number of abnormal behaviors required to characterize a valid diagnosis i.e., identifying the ‘syndromic threshold’ for a given diagnosis. Data-driven clustering approaches such as latent class analysis derive groups that differ in the number of clinical indicators endorsed ^17^ and thus inform attempts to set syndromic thresholds. Such efforts need to be balanced with clinical applicability in real world settings, which require practical decisions such as how to combine clinical indicators from distinct domains (i.e., irritable mood and outbursts). The latter can be achieved by investigating whether domains explain overlapping or distinct aspects of DMDD latent structure and related impairment, thus determining whether “and” or “or” rules should be used to provide a ‘clinical operationalization’ of the diagnosis. Previous research in pre-adolescents and adolescents suggests irritable mood and temper outbursts predict each other over time. However, while each of them are associated with increased risk for disrupted functioning in adolescents ^15^, current criteria require both to be present for a diagnosis to be assigned.

The aim of this study is to evaluate alternative clinical thresholds for the DMDD diagnosis (see Figure 1 for an overview of the analytic strategy and Methods for details). We investigate 3,562 pre-adolescents/early adolescents aged 10-12. First, we used Confirmatory Factor Analysis (CFA) to identify item-level thresholds differentiating normative from clinical problems (the symptomatic threshold). This was used to dichotomize response levels as clinically significant or not. We next used these binary clinical indicators as input to a latent class analysis (LCA) that assigned individuals into clusters with high and low levels of clinical indicators for each domain. This was followed by receiver operating curves (ROC) to detect the number of clinical indicators needed to predict class membership from the LCA and to translate the data-driven results to DSM-5 symptom counts (the syndromic threshold). We then compare the impact of AND/OR rules on impairment and dimensional levels of psychopathology (clinical operationalization). Finally, we investigate the impact of varying definitions on DMDD prevalence and comorbidity profiles in a population-based sample.

**Figure 1.**
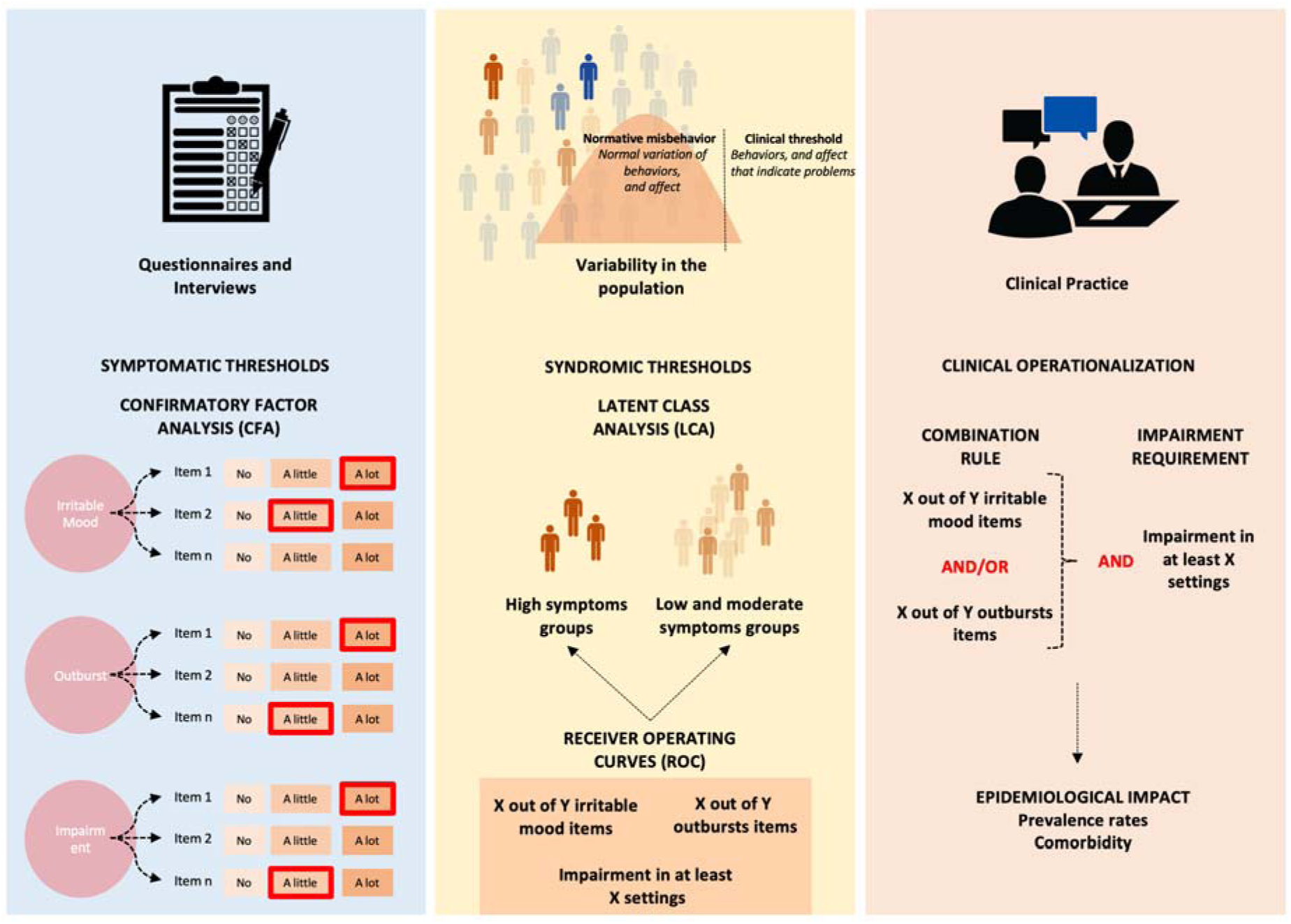
Symptomatic and syndromic thresholds and clinical operationalizations.

## Method

### Participants

Participants of this study were pre-adolescents/early adolescents aged 10-12 from the 2004 Pelotas Birth Cohort Study. All births occurring in the city of Pelotas, from January 1st to December 31st, 2004 were enrolled and followed over time. Pelotas is in southern Brazil and has a population of 328,000. For a full description of the methods, see ^18^. Briefly, all 4231 live births in the city in 2004 whose mothers lived in the urban area and agreed to participate in the longitudinal study were considered eligible. Follow-up home visits were performed when the subjects had reached the ages of 3.0 months (SD=0.1), 11.9 months (SD=0.2), 23.9 months (SD=0.4), and 49.5 months (SD=1.7). When the subjects were, on average, 6.8 years old (SD=0.3) and 11.0 years old (SD=0.4), additional follow-up visits were conducted at a research clinic run by the Postgraduate Program of Epidemiology (Faculty of Medicine, Federal University of Pelotas, Brazil). Of the 4,231 subjects in the original birth cohort, 3,562 (84.1%) were included in our analysis, which used all available data from the 10-12 years of age assessment. The sample comprises 2,353 participants aged 10, 1,206 aged 11 and 4 aged 12. The prevalence of DMDD in this sample using current criteria associated with clinical ratings was 2.5% (95% CI=2.0–3.0) ^19^. The study was approved by the Research Ethics Committee of the Federal University of Pelotas and by the Research Committee of the University of São Paulo School of Medicine. Written informed consent was obtained from all subjects.

### Instruments and Diagnostic Assessment

The parent-version of the DMDD section from the Development and Well-Being Assessment (DAWBA) questionnaire ^20^ was administered by certified psychologists. This questionnaire uses open and closed ended questions to identify the occurrence of clinical indicators in children and adolescents aged 5-17, based on the DSM criteria. The closed ended questions start with two skip questions about the frequency of temper outbursts and irritable mood. Parents who answered that temper outbursts and/or irritable mood occurred at least once a week were probed to answer specific questions that characterize all DSM-5 criteria for DMDD.

A total of 593 parents of participants answered the DMDD section on irritable mood, representing the top 17% of irritable mood frequency. This section includes 9 items characterizing the threshold for experiencing anger, intensity of anger if compared to peers of the same age, duration of anger during the day, whether irritable mood is perceived by others, setting in which anger occurs (at home, at school, with peers) and number of anger weeks throughout the year. A total of 425 parents answered the DMDD section on outbursts, representing the top 12% of frequency of outbursts. This section includes 15 items describing behavior during outbursts (slamming doors, shouting, swearing, saying mean things to others, saying negative things about self, physical aggression to others, deliberate self-harm, breaking things), setting in which outbursts occurred (at home, at school, with peers) and triggers (recognizable and easily triggered). We do not use the item “outbursts free-gap in the last year” in our analysis (DSM requires that there is not a period higher than 3 or more consecutive months without irritable mood and temper outbursts). The rationale for excluding this item is that it is unclear whether we would expect this item to be monotonically related to the overall latent construct given short periods of irritability with large gaps could also inform episodes of irritability (a marker of severity and bipolar disorder).

Lastly, 686 mothers or caregivers that completed either the outburst or irritable mood sections were asked to also complete 4 items about impairment (impact on family life, friendship, learning, and leisure activities). After the impairment questions, mothers or caregivers answered the open-ended questions that allow qualitative description of the symptoms, frequency, and other characteristics of the disorder. All questions and response categories from the DMDD section are depicted in Table S1, available online.

The DAWBA was administered to mothers or caregivers by trained psychologists. The forty-hour training included lectures, role playing, and supervised clinical interviews with pediatric and mental health outpatients at the Federal University of Pelotas. The clinical evaluation of the total sample was performed by a psychologist, and a second independent psychologist evaluated 10% of the study sample. Both were trained in how to apply the DAWBA, in a standardized manner, by the child psychiatrist who had translated and validated the questionnaire for use in Brazil ^21^. Rating procedures were used for assigning comorbidities given DMDD diagnosis was performed *a posteriori*. The inter-rater agreement was 91.2% for the presence of any psychiatric disorder, 75.9% for any anxiety disorder, 73.5% for any depressive disorder, 72.7% for ADHD, 72.9% for conduct disorder, 85.6% for any autism spectrum disorder, 59.5% for any eating disorder, and 52.4% for any tic disorder. Details of the questionnaire can be found online and in other studies ^22^.

The Strengths and Difficulties Questionnaire (SDQ) was used to measure dimensional psychopathology. The SDQ is a 25-item behavioral screening questionnaire with five domains, each of which contain five items (emotional, conduct, hyperkinetic, peer relationships, prosocial behaviors and impact scores). The overall SDQ total scores had a Cronbach’s alpha of 0.82, which is considered high. Internal consistency for the SDQ subscales were low to moderate ranging from 0.48 (peer relationships) to 0.78 (hyperkinetic). Despite low reliability, we maintained results from subscales for their descriptive nature in Supplement 1, available online.

### Statistical Analysis

#### Symptomatic threshold

The fourteen items on outbursts, eight items on irritable mood and four items on impairment were included in three CFAs testing unidimensional models for each construct (n= 593, 425 and 685 respectively). CFA models estimate item level factor loadings (λ) and response category thresholds. Factor loadings represent the strength of the relationship between the latent trait and the item, i.e., they indicate how well each item discriminates different severity levels of a given construct. Category thresholds indicate the expected value of the latent factor at which there is a 50% probability of endorsing a given category or higher i.e., the category threshold indicates the severity level at which the transition from one response category to the next is likely to happen (e.g., from ‘No’ to ‘A little’ or higher, or from ‘A little’ to ‘A lot’).

To distinguish normative misbehavior from behavior that would meet a diagnostic criterion, we used category thresholds from the CFA. CFAs were performed only in subjects with a frequency of irritable mood and outbursts greater than once a week. In this sample, a value of 0.5 represents a half standard deviation above the mean of the distribution of subjects with a frequency of irritable mood and outbursts greater than once a week. Therefore, we interpreted values below 0.5 as typical development (normative) and values at or above 0.5 as ‘clinical indicators’ (a proxy for symptoms or problem indicators). The latter represents an approximation to the top 5% most symptomatic pre-adolescents/early adolescents in the population, which is a threshold used in other diagnostic investigations ^12^. For details about the CFA, see Supplement 1, available online.

#### Syndromic threshold

Before data analysis, each questionnaire item was dichotomized at the value of the category threshold defined in the symptomatic threshold analysis described above in subjects with at least one clinical indicator. Dichotomized items were chosen to enter the LCA analysis because our intention was not to characterize varying levels of irritability in the community, but to identify groups that differ in their number of clinical indicators. Three Latent Class Analyses (LCA) were used to create empirically derived groups with different levels of clinical indicators for irritable mood, outbursts and impairment. Next, we used three Receiving Operating Curves to predict the most accurate number of clinical indicators for detecting participants with high levels of symptoms (as defined by latent class analysis) with regard to irritable mood, outburst and impairment. ROC analysis was used as a way to translate results from the syndromic thresholds of the LCA to the reality of clinical practice, which uses symptom counts. Thus, the ROC identifies a simple rule to allow the identification of patients that are likely to be members of the cluster that exhibit a high level of clinical indicators. The optimal cut-off was estimated using the Younden’s J Statistic, which maximizes both sensitivity and specificity ^23^.

#### Clinical operationalization

Four analytical strategies were used to determine the most appropriate rule for clinical operationalization: the ‘OR’ rule vs. the ‘AND’ rule. First, we compared the fit of CFA models (n=398), putting the selected dichotomized clinical indicators into a unidimensional model of irritability and a correlated model of irritability with two domains (irritable mood and outbursts). Second, we tested whether meeting criteria for the irritable mood group and/or for the outbursts group have distinct or overlapping associations with the impaired functioning group using a multiple logistic regression. Third, we used left censored regressions to compare skewed SDQ dimensional scores between subjects meeting criteria only for irritable mood, only outbursts, either, or both and compared with a group of participants with other DSM disorders except for DMDD and typically developing comparisons. Fourth, for both OR and AND groups, we used a matching procedure to compare levels of SDQ scores between a group that differed in DMDD status (yes vs. no DMDD) but were otherwise fully matched for comorbidities.

#### Epidemiological impact

Finally, using the relative frequency, we investigated the impact of these AND/OR rules and combinations for impairment requirements on the prevalence rates of DMDD in the community and on the comorbidity profile.

All analysis were performed in R version 3.6.1 ^24^, including applications implemented in the packages lavaan 0.6-5 ^25^, poLCA 1.4.1 ^26^, pROC 1.15.3 ^27^, CensReg 0.5-26 ^28^ and MatchIt 3.0.2 ^29^. The R markdown codes for the symptomatic, syndromic and clinical operationalization thresholds of current analysis can be found in Supplement 1, available online.

## Results

### Symptomatic threshold

All eight items of irritable mood were found to be normative in their lowest thresholds and clinical indicators (proxy for symptoms) in the highest thresholds, except “irritable mood that happens at home”, which was found to be normative in all response categories. For the six items that describe intensity, the response option “A little” indicated normative behavior, while the response option “A lot” or “A great deal” indicated a symptom. For the duration item, irritable mood lasting less than an hour indicated normative behavior, whereas irritability lasting a few hours or most of the day indicated a symptom. For the frequency item, irritable mood occurring less than 3 times a week indicated normative behavior, whereas irritable mood occurring every day indicated a symptom (Table S2, available online).

For outbursts, threshold varied substantially across items. For some items, their occurrence even at mild levels indicated a symptom, while other items did not indicate a symptom at any level. Outbursts that include self-harm, breaking things or saying negative things about self, or those that occurred in the classroom are indicative of a symptom if they occur at any level (i.e., “A little” or “A lot”). Outbursts that occurred with peers, include physical aggression, or are easily triggered indicate a symptom when they occurred “A lot”, but were normative when they occurred “A little”. Outbursts that occurred at home and included the pre-adolescents/early adolescents saying mean things, slamming doors, shouting, or swearing did not indicate a symptom irrespective of the level endorsed. Also, whether the triggers were recognizable or not was not relevant to symptom designation. Regarding frequency, only outbursts that occurred daily indicated a symptom (Table S3, available online).

For impairment, “impact on family life” is normative when “A little” and indicate a symptom when “A medium amount” or “A lot”. Impairment that occurs in the other settings (friendship, learning or leisure) indicate a symptom at any level (Table S4, available online). See Figure 2.

**Figure 2.**
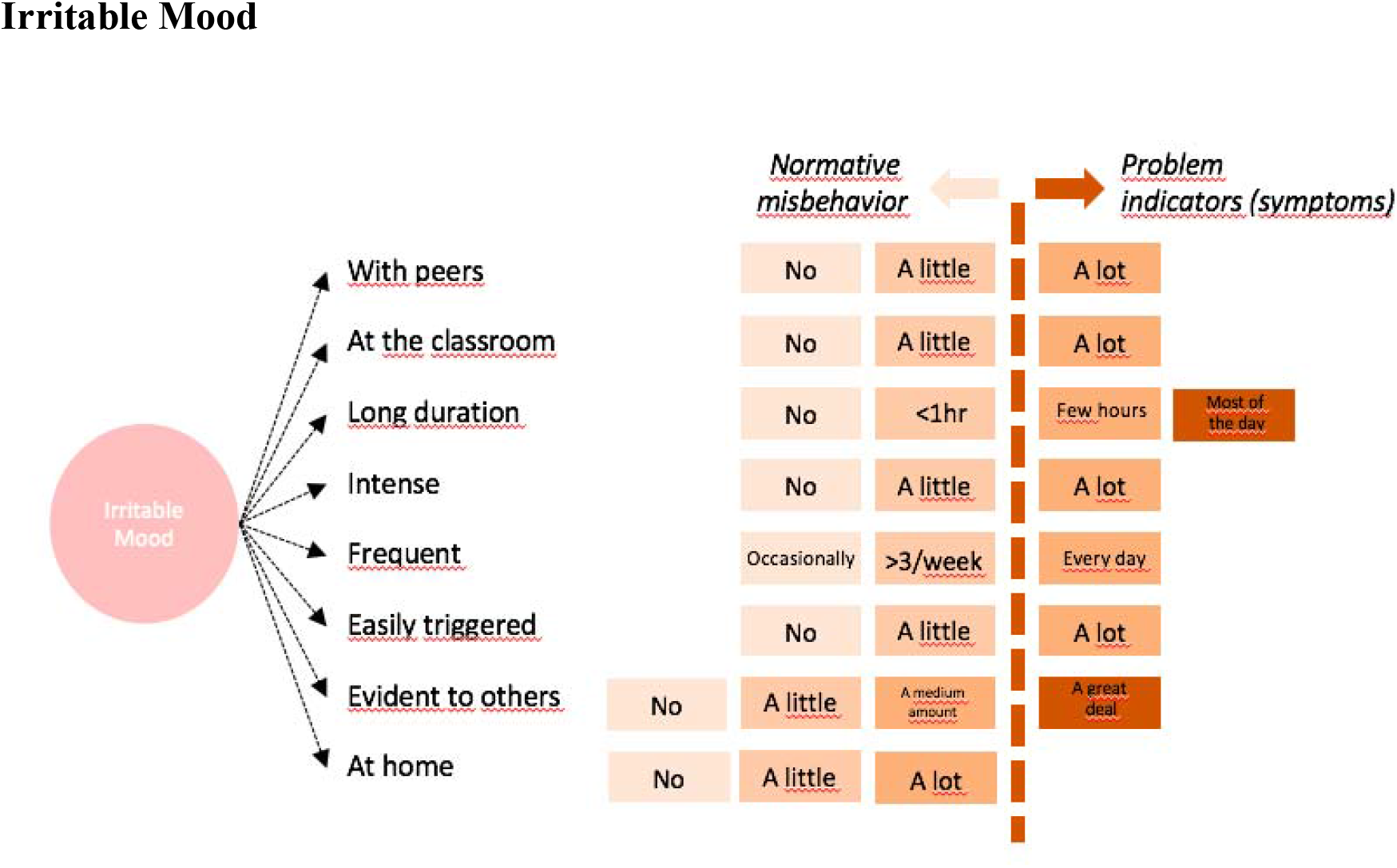
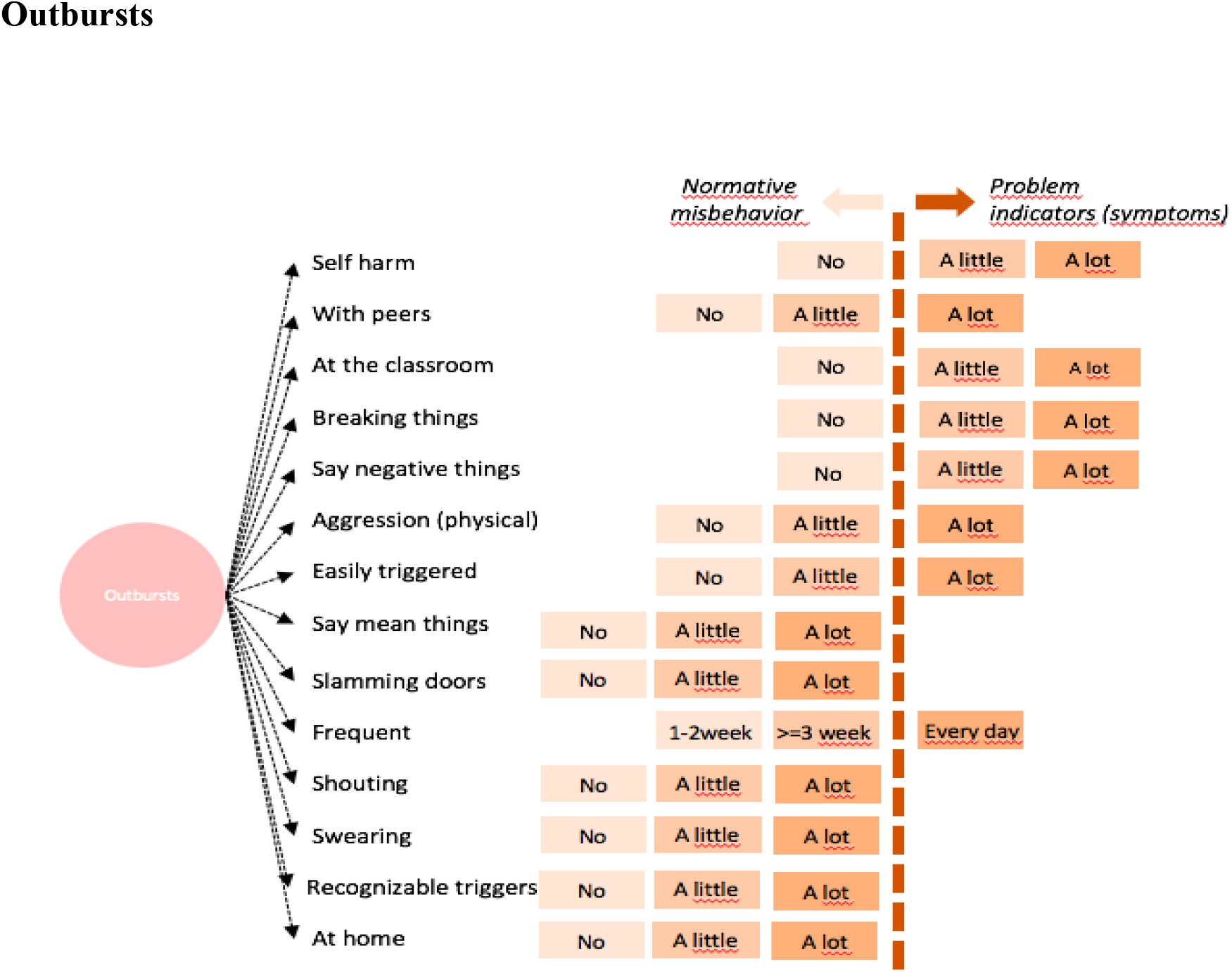
Symptomatic threshold for each irritable mood and outbursts item in the Confirmatory Factor Analysis **Panel A - Irritable Mood** **Panel B - Outbursts**

### Syndromic threshold

The prevalence of each clinical indicator is presented in Table 1. The three LCA analyses (irritable mood, outbursts and impairment) indicated that the two-class solution was the best for each of the three domains (Table S5, available online). This indicates that, in each of the three domains, the population is divided into two groups characterized by high vs. low symptoms (Figure S1, available online). We next performed three Receiver Operating Curve analyses (irritable mood, outburst, impairment) to determine the best number of clinical indicators (i.e., those items identified by the CFA) to use to predict membership in the high vs. low symptom classes identified by the LCA. Younden’s J demonstrates that subjects in the high symptom irritable mood and outburst classes are most accurately characterized by 2 out of 7 irritable mood symptoms and 3 out of 8 outburst symptoms. As for impairment, the subjects with high level of symptoms in LCA are most accurately characterized by significant impairment in at least two settings (Table 2 and Figure S2, available online).

**Table 1.**
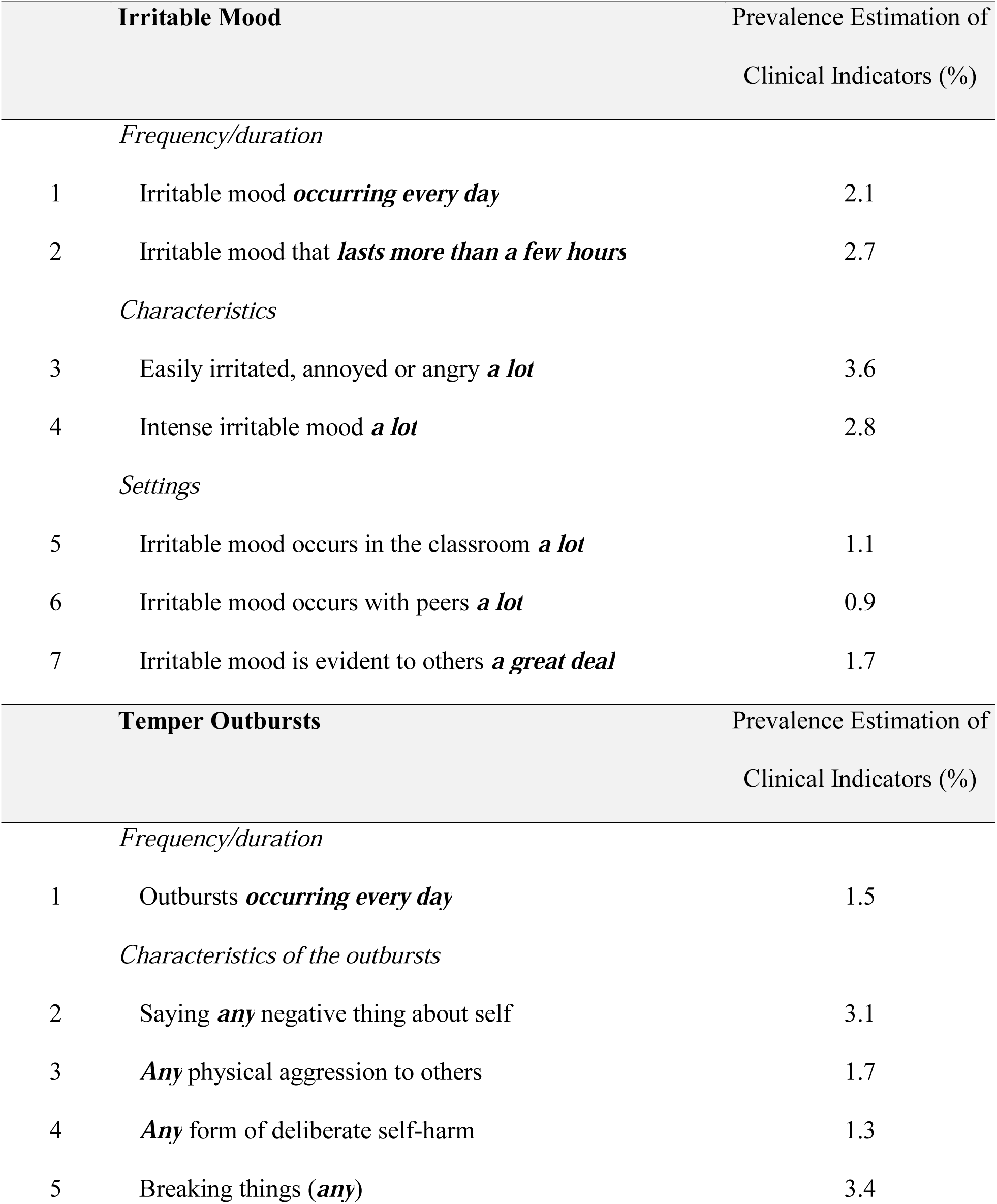
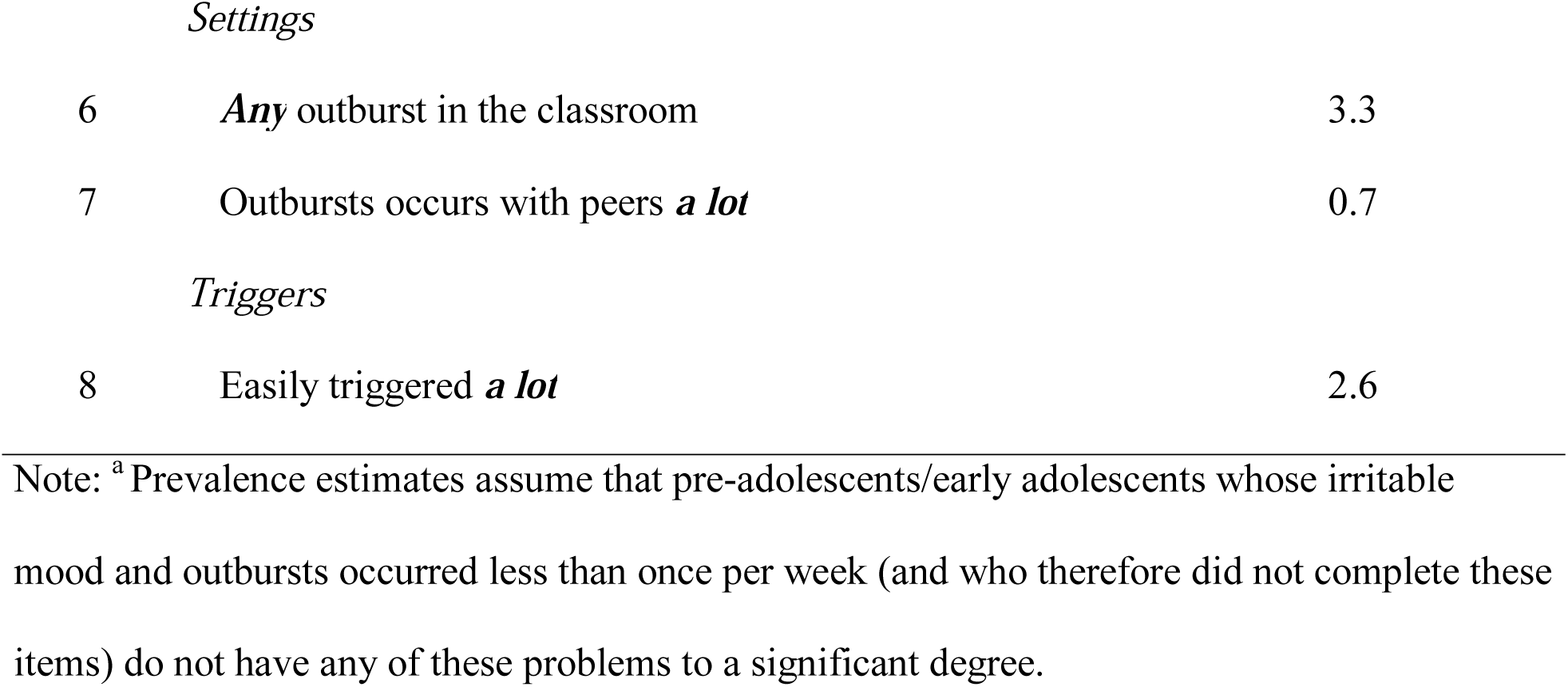
Prevalence^a^ of Each Disruptive Mood Dysregulation Disorder Item Written in Combination With the Response Category That Defines a Clinical Indicator

| **Irritable Mood** |  | Prevalence Estimation of Clinical Indicators (%) |
| --- | --- | --- |
| *Frequency/duration* |  |  |
| 1 | Irritable mood *occurring every day* | 2.1 |
| 2 | Irritable mood that *lasts more than a few hours* | 2.7 |
| *Characteristics* |  |  |
| 3 | Easily irritated, annoyed or angry *a lot* | 3.6 |
| 4 | Intense irritable mood *a lot* | 2.8 |
| *Settings* |  |  |
| 5 | Irritable mood occurs in the classroom *a lot* | 1.1 |
| 6 | Irritable mood occurs with peers *a lot* | 0.9 |
| 7 | Irritable mood is evident to others *a great deal* | 1.7 |
| **Temper Outbursts** |  | Prevalence Estimation of Clinical Indicators (%) |
| *Frequency/duration* |  |  |
| 1 | Outbursts *occurring every day* | 1.5 |
| *Characteristics of the outbursts* |  |  |
| 2 | Saying *any* negative thing about self | 3.1 |
| 3 | *Any* physical aggression to others | 1.7 |
| 4 | *Any* form of deliberate self-harm | 1.3 |
| 5 | Breaking things (*any*) | 3.4 |
| *Settings* |  |  |
| 6 | ***Any*** outburst in the classroom | 3.3 |
| 7 | Outbursts occurs with peers ***a lot*** | 0.7 |
| *Triggers* |  |  |
| 8 | Easily triggered ***a lot*** | 2.6 |

Note: <sup>a</sup> Prevalence estimates assume that pre-adolescents/early adolescents whose irritable mood and outbursts occurred less than once per week (and who therefore did not complete these items) do not have any of these problems to a significant degree.

**Table 2.** Receiver Operating Curves Parameters Investigating the Best Number of Clinical Indicators to Capture Latent Class Groups

| Threshold | Prediction of Latent Class Groups |  |  |  |  |  |  |  |  |  |  |  |  |  |  |  |  |  |
| --- | --- | --- | --- | --- | --- | --- | --- | --- | --- | --- | --- | --- | --- | --- | --- | --- | --- | --- |
|  | Irritable Mood |  |  |  |  |  | Outbursts |  |  |  |  |  | Severity of Impairment |  |  |  |  |  |
|  | ACC | Sens | Spe | PPV | NPV | YI | ACC | Sens | Spe | PPV | NPV | YI | ACC | Sens | Spe | PPV | NPV | YI |
| 0 | 0.25 | 1.00 | 0.00 | 0.25 | - | 0 | 0.12 | 1.00 | 0.00 | 0.12 | - | 0 | 0.36 | 1.00 | 0.00 | 0.12 | - | 0 |
| 1 | 0.56 | 1.00 | 0.41 | 0.36 | 1.00 | 0.4 | 0.40 | 1.00 | 0.32 | 0.17 | 1.00 | 0.32 | 0.73 | 1.00 | 0.58 | 0.57 | 1.00 | 0.58 |
| **2** | **0.86** | **0.95** | **0.83** | **0.65** | **0.98** | **0.78** | 0.69 | 1.00 | 0.64 | 0.28 | 1.00 | 0.64 | **0.93** | **1.00** | **0.89** | **0.83** | **1.00** | **0.89** |
| **3** | 0.90 | 0.64 | 0.99 | 0.95 | 0.89 | 0.63 | **0.90** | **1.00** | **0.88** | **0.54** | **1.00** | **0.88** | 0.90 | 0.72 | 1.00 | 1.00 | 0.87 | 0.87 |
| 4 | 0.83 | 0.32 | 1.00 | 1.00 | 0.82 | 0.32 | 0.98 | 0.86 | 0.99 | 0.98 | 0.98 | 0.85 | 0.74 | 0.28 | 1.00 | 1.00 | 0.71 | 0.28 |
| 5 | 0.78 | 0.11 | 1.00 | 1.00 | 0.77 | 0.11 | 0.93 | 0.43 | 1.00 | 1.00 | 0.92 | 0.43 |  |  |  |  |  |  |
| 6 | 0.76 | 0.04 | 1.00 | 1.00 | 0.76 | 0.04 | 0.89 | 0.14 | 1.00 | 1.00 | 0.89 | 0.14 |  |  |  |  |  |  |
| 7 | 0.75 | 0.00 | 1.00 | - | 0.75 | 0 | 0.89 | 0.12 | 1.00 | 1.00 | 0.89 | 0.12 |  |  |  |  |  |  |
| 8 |  |  |  |  |  |  | 0.88 | 0.02 | 1.00 | 1.00 | 0.88 | 0.02 |  |  |  |  |  |  |

Note: ACC = Accuracy; NPV = Negative Predictive Value; PPV = Positive Predictive Value; Sens = Sensitivity; Spe = Specificity; YI = Youden's Index.

### Clinical operationalization

First, a model with two correlated domains (irritable mood and outbursts) provided a better fit than a unidimensional model encompassing both domains (χ^2^_diff_=7.3, df=1, p=0.007; Table S6, available online). Second, both irritable mood and outbursts were associated with clinical impairment in univariate models (irritable mood OR=41.71, p<0.001; outbursts OR=76.1, p<0.001) and in multiple models adjusted for the effects of including both predictors in the same model (irritable mood adjusted OR=18.2, p<0.001; outbursts adjusted OR=23.63, p<0.001). Third, comparisons between irritable only, outbursts only and combined groups with typically developing comparisons and with a group of patients with other DSM disorders (except for DMDD) showed all three DMDD groups had higher scores on all SDQ scales than typically developing comparisons and higher total SDQ total scores than subjects with other DSM diagnosis (Figure S3; Table S7, both available online). Fourth, left-censored regressions comparing groups matched for comorbidity (any anxiety, any mood, any hyperkinetic and any disruptive behavior disorder) showed that, using either the OR or the AND rule, the DMDD group showed higher total, emotional, conduct, hyperactivity, peer relationship and impact scores than did the non-DMDD group with matched comorbidities (Figure S4 and Figure S5, available online).

### Epidemiological impact

When using an “OR” rule, the optimal criteria from the ROC analysis (2 of 7 irritable mood symptoms, 3 of 8 outburst symptoms, impairment in at least two settings) resulted in a prevalence of 3.0%: 1.12% have only irritable mood, 0.64% only outbursts and 1.23% have both irritable mood and outbursts (Table 3). Both the “OR rule” and the “AND rule” resulted in higher levels of psychiatric comorbidities compared to the current DMDD clinical criteria (Table S8, available online).

**Table 3.** Impact of Different Rules for Combining Irritable Mood and Temper Outburst Clinical Indicators and Impairment Requirements on Prevalence Rates of Disruptive Mood Dysregulation Disorder (DMDD)

|  | **Irritable**<br>**Mood** | **Outbursts** | **AND Rule** | **OR Rule** |
| --- | --- | --- | --- | --- |
| No impairment requirement | 2.41 | 0.9 | 1.63 | 4.94 |
| At least one setting | 1.80 | 0.70 | 1.46 | 3.96 |
| **At least two settings (optimal)** | **1.12** | **0.64** | **1.23** | **3.00** |
| At least three settings | 0.61 | 0.48 | 0.87 | 1.96 |
| All four settings | 0.17 | 0.17 | 0.53 | 0.87 |

Note: Settings: 1= Impact on family life; 2 = impact on friendship; 3 = Impact on learning; 4 = Impact on leisure.

## Discussion

This study provides important information to guide a revision of the diagnostic criteria for DMDD. Using CFA, we found that seven of the eight irritable mood items were normative when endorsed in the low response categories and clinical indicators in the high response categories. The one exception was “irritable mood that happens at home”, which was always normative. For outbursts, the threshold for a clinical indicator varied substantially across items. For some items, such as outbursts with self-harm, their presence indicated a problem even at only mild levels. Others, such as shouting, were not clinical indicators even when present at the highest threshold. ROC analyses indicated that a combination of 2 of 7 irritable mood symptoms, 3 of 8 outburst symptoms and significant impairment in at least two settings would best predict membership in the “high” vs. “low” LCA-based symptom classes. The four clinical operationalization analysis converge to demonstrate that the two domains differ from a latent perspective; they are independently associated with impairment; and OR-rule groups show comparable or even higher levels of impairment then other DSM disorders. Matched analysis showed that results cannot be attributed to comorbidity. The most accurate solution resulted in a prevalence of 3% in the fully automated operationalized criteria (1.12% only irritable mood, 0.64% only outbursts and 1.23% combined).

Our findings are consistent with the limited literature examining irritability dimensionally in the population. Each set of findings suggest that normative outbursts differ from clinical indicators in frequency, duration, quality, context, and triggering events ^30–33^. Wakschlag et al. ^12^ found that outbursts characterized by high frequency, “long duration”, or “aggressive components”, or those that occurred “with nonparental adults” or “out of the blue” were clinical indicators. Wiggins and collaborators ^13^ also used an empirical approach to identify irritable behaviors indicative of problems in preschoolers.

They examined 22 temper loss behaviors from the criteria for oppositional defiant disorder, DMDD and other depressive disorders in the DSM-5 and found two informative items. Similar to our work, the item “easily frustrated” indicated a symptom only when present nearly every day, but the item “break/destroy” indicated a symptom even when at lower frequencies. Nevertheless, those thresholds might vary substantially in distinct age ranges and cultures, which highlights the need for developmentally sensitive studies.

Clinical operationalization analysis suggests that an OR rule is most appropriate to capture cases in need of treatment. This algorithm identified pre-adolescents/early adolescents with either irritable mood or outbursts who manifested associated impairment, elevated symptoms, and functional impairment. This resulted in a prevalence rate of 3%, which is higher than the prevalence rate of 2.5% by the current diagnostic criteria. Of course, it is not possible to identify the “true” prevalence of DMDD in the population with one study; rather the current analyses inform nosologists’ attempts to weigh the strengths and weaknesses of various diagnostic thresholds.

Advancing understanding about DMDD diagnostic criteria is a major concern in children and adolescent psychiatric practice. Our findings are a first step towards defining parameters to alert the clinician when to be (and when not to be) concerned with irritable mood and outbursts. Our approach suggests several refinements to the DSM-5 criteria. First, the new criteria provide a list of behaviors and a threshold for each that specifies when to consider that behavior to be a clinical indicator. This is more descriptive, precise, and data-based than the current criteria and provide a way ‘calibrate’ the severity of each clinical indicator composing the syndrome. Second, we suggest a syndromic threshold for the combination of such behaviors. This is a more practical way to separate normal from abnormal behaviors and considers that DMDD might present itself with distinct clinical indicators. Third, our data support an OR rule when combining irritable mood and outbursts, rather than the AND rule currently in the manual. Finally, our results support the importance of requiring two settings for the diagnosis, as in DSM-5. Specifically, our data indicate that, while the impact of symptoms on function needs to be at medium levels on family life to be considered a clinical indicator, mild levels of impairment in friendship, learning or during leisure activities should suffice as a clinical indicator for the DMDD impairment criteria.

Our study has important strengths. *First*, we relied on a large representative population sample and implemented assessment methods that could mimic clinical assessment in the real world, as far as possible in an epidemiological investigation. *Second*, we applied Confirmatory Factor Analysis, Latent Class Analysis and Receiver Operating Curves Analysis, applying a similar framework used in other disorders ^36,37^ to a new syndrome that lacks empirical investigations to guide operationalization. However, this work has also some important limitations. First, our analysis is focused on internal validators. Further studies investigating course, family history, treatment response, and other external validators are needed to demonstrate the validity of the operationalized syndrome. Since associations between symptoms and irritability-related impairment were investigated using the same DAWBA DMDD section, the size of the associations is likely to be overestimated. However, the value of these odds ratios may be helpful in understanding whether the two aspects of irritability capture distinct or overlapping aspects of irritability-related impairment. Second, our subjects were all 10-12 years old, and our data might not be generalized to other developmental stages. Third, because of the skip rule questions, the CFA parameters were estimated for subjects with irritable mood or outbursts that occurred at least once a week. Analysis were modeled to consider these characteristics, but this might have biased the parameter estimates for some items. Also, our approach assumes irritable mood, outbursts and impairment are distinct domains, which is still an empirical question to be further tested. Fourth, our analysis is restricted to parent reports, and no information was acquired from pre-adolescents/early adolescents themselves. Lastly, our approach is restricted to a single sample and it is unclear whether those results can be replicated in other samples.

To conclude, this is the first study in the field with this intent in this age range and thus is a first step towards refining the diagnostic criteria of DMDD. Future research to advance the field of DMDD should include replicating these findings; extending similar approaches to diagnostic instruments other than the DAWBA; examining symptomatic thresholds using measures that do not have skipping rules and are designed specifically to differentiate normative versus non-normative behaviors; investigating interrater reliability; and including developmentally sensitive items and external validators. Furthermore, prospective longitudinal investigation that applies this framework beginning at earlier ages can elucidate the origins of pathologic irritability, thus guiding the development of novel interventions and developmentally-based prevention.

## Data Availability

Available upon request to the Pelotas Graduate Program.

## Data Availability

Available upon request to the Pelotas Graduate Program.

